# Analysis of the diabetic arterial transcriptome to define novel biomarkers of macrovascular disease

**DOI:** 10.64898/2026.02.08.26345847

**Authors:** Aya Shouma, Marilena Giannoudi, Marcella Conning-Rowland, Michael Drozd, Oliver I Brown, Chew Cheng, Piruthivi Sukumar, Katherine I Bridge, Eylem Levelt, Marc A Bailey, Kathryn J Griffin, Mark T Kearney, Richard M Cubbon

## Abstract

**Objective:** Diabetes mellitus (DM) approximately doubles the risk of atherosclerotic cardiovascular disease (ASCVD) events, but the molecular basis is poorly understood. We aimed to define arterial differentially expressed genes (DEGs) associated with DM, validate hits as plasma proteins, and ascertain whether these complement ASCVD risk prediction tools.

**Research design and methods:** RNA-sequencing data from the Genotype-Tissue Expression (GTEx) cohort was used to define DEGs associated with DM in two arterial sites in >90 people with DM and >330 controls. UK Biobank (UKB) was used to corroborate that DEGs in their plasma protein form were differentially abundant in people with DM and associated with ASCVD events. Finally, we assessed if including these plasma proteins improved performance of the SCORE2 and SCORE2-Diabetes ASCVD risk models.

**Results:** 619 and 356 DEGs were associated with DM in the thoracic aorta and tibial artery, respectively. Of these, 22 were common to both arteries, all of which were directionally concordant. Of these, 5 were included in the UKB plasma proteomics dataset and we corroborated 4 (ACP5, LEFTY2, LILRA5 and PSME2) as showing concordant differential abundance in people with DM; all demonstrated associations with a range of incident ASCVD events. Addition of the 4 proteins to SCORE2 and SCORE2-Diabetes (for people without and with DM, respectively) improved the population-level discrimination, classification and calibration of these models.

**Conclusions:** DM is associated with a distinct arterial gene expression profile, hits from which are associated with ASCVD events and add value to risk prediction.

**Visual abstract:** 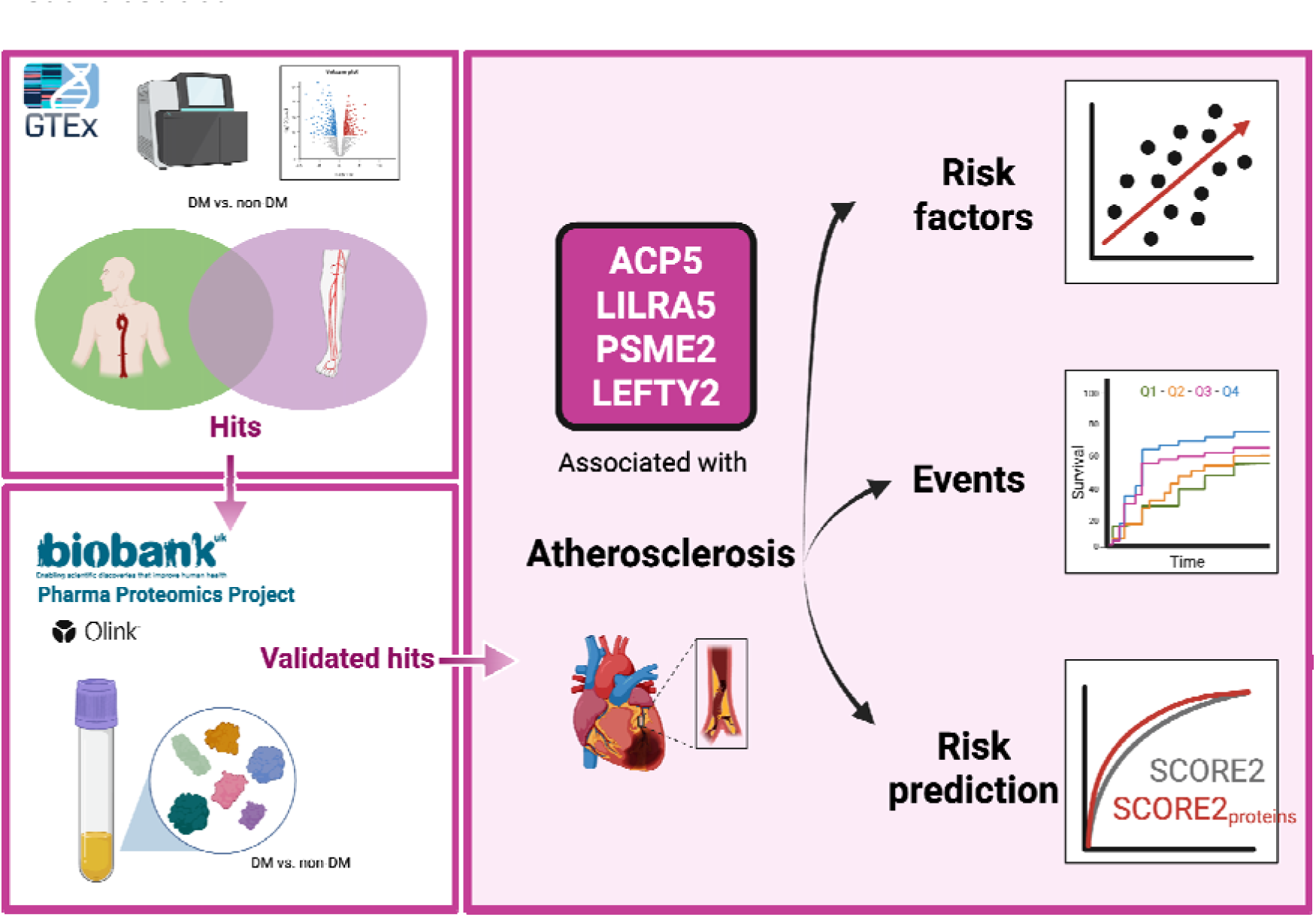

## Introduction

Cardiovascular disease (CVD) remains one of the leading causes of morbidity and mortality worldwide, with more than 20 million deaths attributed in 2021. Atherosclerotic CVD (ASCVD) accounts for 85% of these deaths (1). Diabetes mellitus (DM) approximately doubles the risk of ASCVD, meaning over half of deaths in this population are attributable to ASCVD (2). The global prevalence of DM among adults aged 20-79 was estimated at 11.1% (589 million) in 2025 and is expected to increase to 853 million by 2050 (3). Therefore, the issue of DM-associated ASCVD poses a major threat to individuals and society in the coming decades.

Coronary computed tomography angiography (CCTA) data show that people with DM experience more widespread and severe coronary atherosclerosis, along with greater arterial calcification (4). Invasive optical coherence tomography (OCT) imaging has corroborated this calcification and revealed a greater volume of lipid in the arterial wall (5). At a histological level, DM is associated with greater macrophage infiltration, more healed plaque ruptures and more abundant calcification of the coronary arterial wall (6). Notably, DM predisposes to atherosclerosis across vascular beds, with analysis of the multi-ethnic study of atherosclerosis cohort showing increased prevalence of coronary, carotid, aortic and peripheral arterial disease (7).

Whilst the cellular and molecular basis of atherosclerosis has been extensively studied, especially in preclinical models, our understanding of how DM impacts the human arterial wall is limited. With the advent of ‘omics technologies, it is now possible to gain unbiased molecular profiles that inform mechanistic understanding and biomarker discovery. We hypothesized that gene expression (transcriptomic) differences associated with DM in the aorta and tibial artery would define the basis of diabetic arterial disease and that common hits could guide selection of plasma protein biomarkers which add value to existing clinical risk models.

## Methods

### Arterial RNA-sequencing data from the GTEx project

The Genotype-Tissue Expression (GTEx) project was established by the National Institutes of Health (NIH) and Broad Institute of MIT and Harvard in 2010 to provide a comprehensive repository of human multi-omic data. This includes RNA-sequencing from 54 tissues (8,9), collected post-mortem from organ donors with family consent, as described in detail on the GTEx project website (https://www.gtexportal.org/). Thoracic aorta transcriptomes are available from 472 donors, of which we included 427 (91 DM vs. 336 non-DM) after excluding those with missing metadata required for covariates in RNA-seq analysis. Similarly, tibial artery transcriptomes are available from 691 donors, of which we included 645 (160 DM vs. 485 non-DM). Bulk RNA-sequencing raw count data were downloaded directly from the GTEx portal, using GTEx version 8 (released 18^th^ July 2019). This includes metadata on donor age, gender, ischaemic time (between death and sample collection), and Hardy score (mode of death classification). Access to sensitive metadata related to comorbidities was obtained through an approved controlled-access application via dbGaP (https://www.ncbi.nlm.nih.gov/gap/) #32524 ‘Defining tissue-specific transcriptional profiles associated with diabetes’.

### RNA-sequencing analysis

Differential gene expression analysis was conducted using R (Version 4.1.1) and the DESeq2 package (Version 1.36.0), using a pipeline we have previously published (10,11). We sought differentially expressed genes (DEGs) associated with DM in separate analyses of thoracic aorta and tibial artery. Donors with type 1 or type 2 DM were pooled to create a single DM variable, since multiple donors were recorded as having both forms of DM and detailed treatment data were not available. Those with DM status unknown were excluded. Covariates in DESeq2 analyses were age (20–29, 30–39, 40–49, 50–59, 60–69, and 70–79); sex; race; ischaemic time as categories of 300 min (0–299, 300–599, 600–899, 900–1199, and 1200–1499); Hardy score (a mode of death classification); BMI which was classed as normal weight (18.5–24.9 kg/m^2^), overweight (25–29.9 kg/m^2^), and obese (>30 kg/m^2^); medical history indication of hypertension and myocardial infarction (MI); and RNA integrity score (RIN; 5.1–6, 6.1–7, 7.1–8, 8.1–9, and 9.1–10). Technical factors including ischaemic time, defined by GTEx as ‘time from death or withdrawal of life-support until the time the sample is placed in a fixative solution or frozen’, Hardy score, and RIN were included. Biological covariates of age, sex, race, and BMI were included because these are also known to alter gene expression in many tissues. Genes were filtered to include only those with greater than 10 read counts in at least the same number of samples as included in the DM subgroup. Effect size shrinkage using the apeglm method was applied for visualization and ranking of genes as this reduces false positive hits in weakly expressed genes (10). False discovery rate (FDR)–adjusted P values produced by DESeq2 using the Benjamini–Hochberg method were used, with adjusted P<0.05 defined as statistically significant. To define biological themes within DEGs, g:Profiler (https://biit.cs.ut.ee/gprofiler/gost) was used to perform enrichment analysis amongst the Gene Ontology (GO) term ‘Biological Process’ and ‘Molecular Function’ gene sets (12). Statistical significance was determined using Benjamini–Hochberg False Discovery Rate (FDR) to account for multiple comparisons, with significance defined as FDR<0.05.

### UK Biobank study population

UK Biobank (UKB) is a large-scale, prospective cohort study that recruited 503,317 individuals aged between 37 and 73 from 22 assessment centres across the UK between 2006 and 2010. It provides detailed multi-omic, imaging and health-related data, accessible via application (as described at https://www.ukbiobank.ac.uk/). It received ethical clearance from the National Health Service Research Ethics Service (Ref: 11/NW/0382), all participants gave written informed consent, and the study was performed in line with the Declaration of Helsinki. All analyses were performed via the secure UK Biobank Research Analysis Platform under application number 528243.

### Definition of Diabetes and Covariates in UKB

Details of UKB data fields and data codes used to define DM and all covariates are shown in **Supplementary Table 1.** DM status was ascertained from participants during a nurse-led interview during their recruitment visit (date defined using UKB Data-Field ID 53). Physical assessments, clinical blood assays, and other covariate data were obtained at the time of recruitment. Estimated Glomerular Filtration Rate (eGFR) was calculated using the R package transplantR v0.2.0 (https://cran.r-project.org/web/packages/transplantr/index.html). As we have previously described (13), pulse-wave arterial stiffness index (PASI) was measured at recruitment in a subset of 162,029 participants. Carotid artery ultrasound was performed in a subset of 41,442 participants as part of a multimodality imaging assessment starting from 2014, when all surviving participants were invited by e-mail and then postal mail. As we have previously described (13), carotid intima-media thickness (CIMT) was assessed at two angles for each common carotid, giving a total of four CIMT measurements, from which mean CIMT was calculated.

### Definition of outcome measures in UKB

In the first phase of UKB analyses exploring associations of plasma proteins with outcomes, we defined the primary outcomes as adjudicated fatal or non-fatal ASCVD events, including non-fatal MI, non-fatal ischemic stroke, and cardiovascular mortality. All-cause mortality was also studied for reference. Cardiovascular events were identified using dates of ST-segment Elevation Myocardial Infarction (STEMI, UKB field ID 42002), non ST-segment Elevation Myocardial Infarction (NSTEMI, UKB field ID 42004), and ischemic stroke (UKB field ID 42008) using a censoring date of 31^st^ October 2022. Mortality outcomes were obtained from official national death registries: NHS Digital for England and Wales, and the NHS Central Register for Scotland. Cardiovascular mortality was defined using ICD-10 codes I00–I99, and all-cause mortality was captured via date of death up to the censoring date of 31^st^ May 2024. Time-to-event analyses commenced at the baseline visit to the UK Biobank assessment centre (UKB field ID 53) and continued until the first occurrence of a cardiovascular event or censoring date.

In risk prediction analyses, we refined the outcome definition to align with the SCORE2 and SCORE2-Diabetes models, focusing specifically on predicting 10-year risk of first fatal or non-fatal ASCVD event (14). For this part, ASCVD events were identified using validated ICD-10 codes (**Supplementary Table 2**), consistent with the methodology outlined by the model derivation paper (14). The follow-up period was limited to 10 years from baseline. This distinction allowed for model evaluation and comparison under the same assumptions used in the original SCORE2 framework.

### Plasma proteomic data in UKB

UKB includes plasma proteomic profiles from 54,219 UKB participants, measured with the Olink Explore 3072 panel, a proximity extension assay using paired antibodies and complimentary oligonucleotides, to quantify 2,923 unique proteins. Details of quality control procedures for plasma proteomics have been descried by Sun and colleagues (15). Protein concentration is provided as normalized protein expression (NPX) values. Where proteins were below the limit of assay detection in specific samples, these are recorded as missing data. We used data collected at the baseline study visit, not the repeat assay data collected in a subset of participants.

### Statistical Analysis

Analyses were performed in Posit Workbench RStudio version 2.2.1 (R version 4.4.0) within the UK Biobank Research Analysis Platform (UKB-RAP). All statistical tests were two-sided, and statistical significance was defined as p<0.05. Missing data were not imputed. All continuous variables were found to be non-normally distributed using the Shapiro Wilk test, so are presented as median with 25^th^–75^th^ centiles. Categorical data are presented as counts with percentages. Where counts are below 50, we do not specify numbers (i.e. small number suppression) to reduce the risk of unintended participant deanonymisation. Comparison of continuous data between groups used Mann Whitney tests, whilst categorical data were compared with Chi-squared tests. Association of plasma proteins with cardiovascular imaging phenotypes used linear regression. For survival analyses, Kaplan Meier curves and log-rank tests were used to compare unadjusted event rates across quartiles of plasma proteins, with Cox proportional hazards regression models used to quantify unadjusted and adjusted hazard ratios. Where indicated, some analyses were repeated after stratification by DM status or with the inclusion of interaction terms to quantify differential associations across DM strata.

When ascertaining the incremental value of plasma proteins in 10-year ASCVD risk prediction, the SCORE2 model covariates were first refitted using data from all UK Biobank participants without DM to assure optimal model performance. This process was repeated for SCORE2-Diabetes using data from all individuals with DM. Logistic regression modelling was performed with a binary 10-year ASCVD event outcome using the R function ‘glm’. The base model was refitted SCORE2 (or SCORE2-Diabetes), represented as all SCORE2 (or SCORE2-Diabetes) risk factors as model covariates. Model outputs were generated with the R function ‘predict’. Extended models used the same base model plus protein biomarkers of interest, included as continuous variables in NPX units. To define if the protein-extended models exhibited superior performance to base models, a range of metrics were assessed. Goodness of fit was compared with likelihood ratio tests using ANOVA function. Model calibration was also assessed by plotting observed event rates against expected rates in deciles of predicted probability, with the 45° line (i.e. y=x) representing perfect calibration. The Expected/Observed (E/O) event ratio was calculated as the mean expected probability divided by the mean observed event rate. To assess the incremental discriminative performance of protein-extended models, receiver operating characteristic (ROC) analyses were performed using the R package pROC v1.19.0.1 (https://cran.r-project.org/web/packages/pROC/pROC.pdf) to calculate the area under the curve (AUC), which were statistically compared using DeLong’s test and plotted using ggplot2 v4.0.0 (https://cran.r-project.org/web/packages/ggplot2/index.html). The continuous Net Reclassification Improvement (NRI) metric was used to assess overall reclassification performance, calculated using the R package nricens v1.6 (https://www.rdocumentation.org/packages/nricens/versions/1.6/topics/nribin). To assess reclassification based on the pre-defined clinical SCORE2 10-year ASCVD risk categories (Low <2%, moderate 2-10%, high 10-20% and very high ≥ 20%), categorical NRI was applied. NRI was calculated with 1,000 bootstrap samples to obtain 95% confidence intervals and NRI scatter plots with marginal density plots were conducted.

## Results

### Defining DEGs associated with DM in the thoracic aorta and tibial artery

Characteristics of participants in the GTEx study with available thoracic aorta and tibial artery data are presented in **Supplementary Table 3**. For both arteries, participants with DM were approximately 7 years older and had approximately doubled prevalence of hypertension and prior MI. After accounting for multiple covariates described in the methods, we identified 619 and 356 DEGs associated with DM in the thoracic aorta and tibial artery, respectively (**Figure 1A-B, Supplementary Tables 4-5**). In thoracic aorta, the 10 most statistically significant enriched GO:BP terms included ‘dendritic cell differentiation’ and ‘response to cytokine’ (**Figure 1C**, **Supplementary Table 6**). In tibial artery, the 10 most statistically significant enriched GO:BP terms included ‘negative regulation of viral process’, ‘regulation of interferon-beta production’ and ‘interleukin-8-mediated signaling pathway’ (**Figure 1D**, **Supplementary Table 7**). At the level of individual genes, we observed 22 common DEGs in thoracic aorta and tibial artery, all of which were directionally concordant, with 8 upregulated and 14 downregulated in the context of DM (**Table 1**). Of the upregulated DEGs, 4 (50%) are annotated by the Human Protein Atlas as belonging to ‘single cell type expression clusters’ attributed to myeloid cells; of the downregulated DEGs, there was more diversity, with fibroblasts being most common in 3 (21%). These data implicate inflammatory and immune factors as an important theme associated with DM in two distinct arterial beds. To validate and extend our observations, we next focussed on shared DEGs with plasma proteomic data available in UKB.

**Figure 1:**
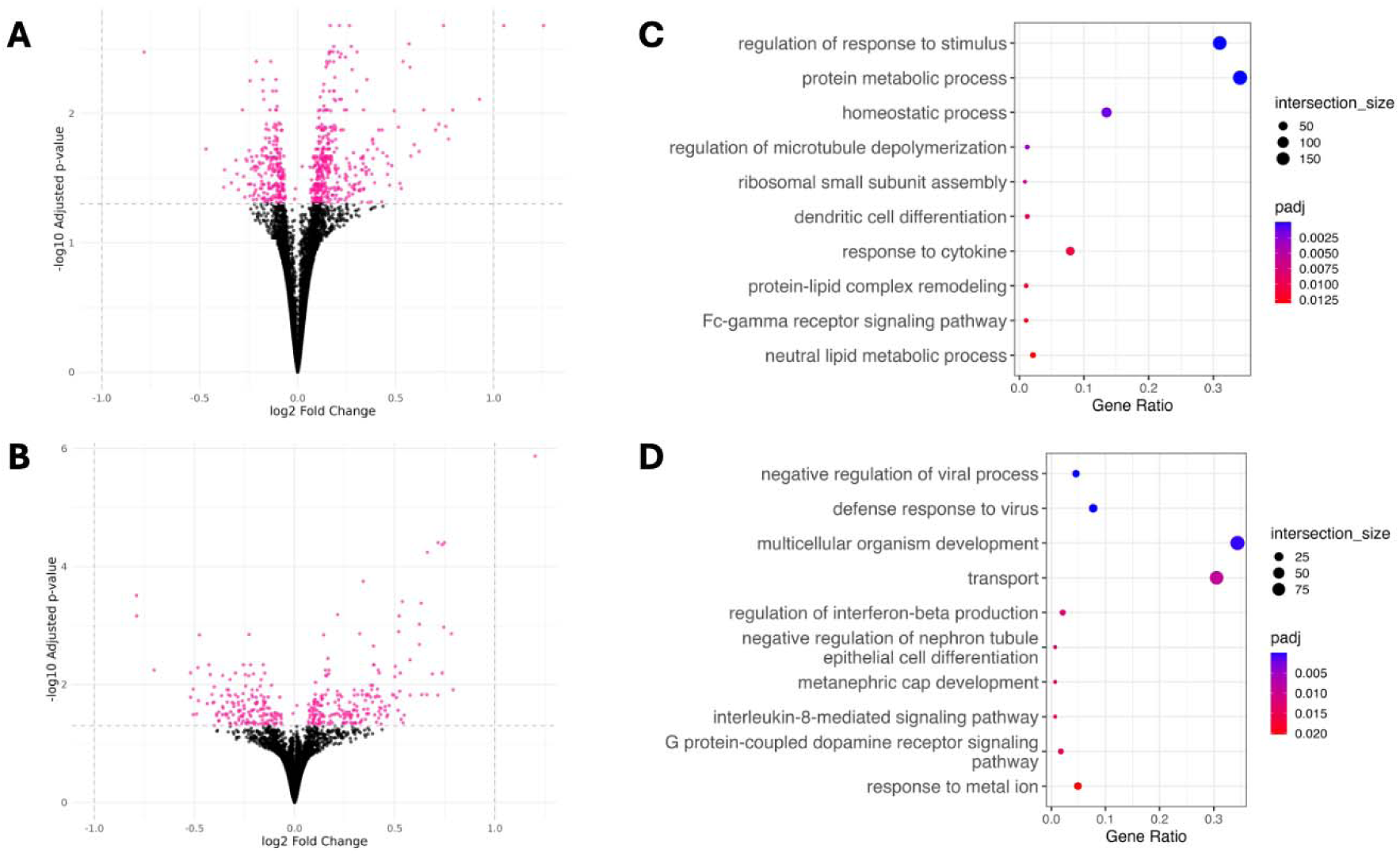
DM is associated with altered gene expression and over-representation of inflammatory gene sets in the thoracic aorta and tibial artery. **A**) Volcano plot illustrating differential gene expression in DM vs. non-DM in thoracic aorta (91 DM vs. 336 non-DM donors). **B**) Volcano plot illustrating differential gene expression in DM vs. non-DM in tibial artery (160 DM vs. 485 non-DM donors). For A and B, each dot represents a gene, with pink colour denoting those that achieve Benjamini–Hochberg FDR-adjusted p<0.05. Raw data are presented in Supplementary Tables 4-5. **C**) Top 10 Gene Ontology Biological Process (GO:BP) terms from thoracic aorta differentially expressed genes (DEGs) in panel A. **D**) Top 10 GO:BP terms relating to tibial artery DEGs in panel B.

**Table 1:**
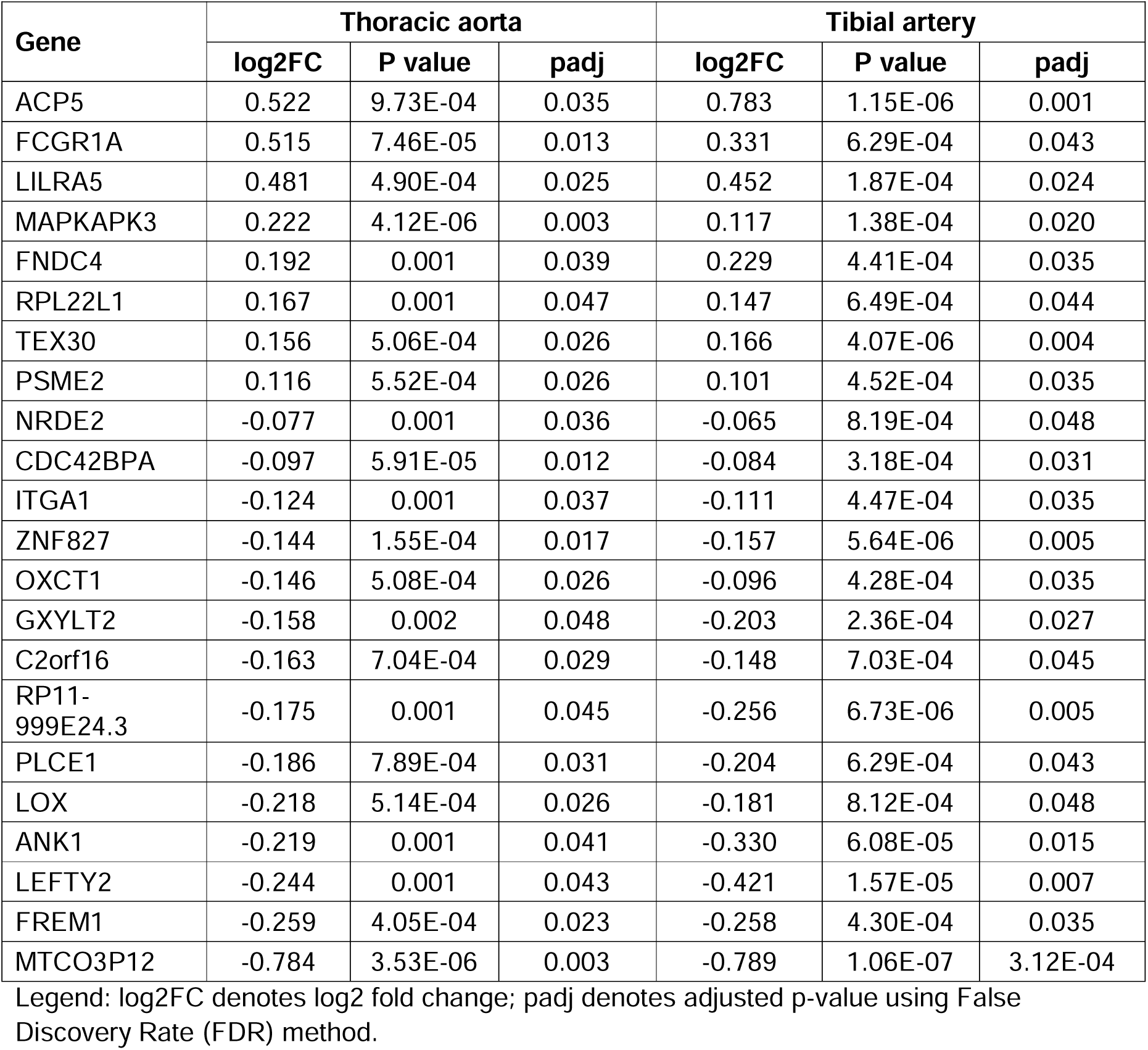
Diabetes-associated differentially expressed genes shared by thoracic aorta and tibial artery.

| Gene | Thoracic aorta |  |  | Tibial artery |  |  |
| --- | --- | --- | --- | --- | --- | --- |
|  | log2FC | P value | padj | log2FC | P value | padj |
| ACP5 | 0.522 | 9.73E-04 | 0.035 | 0.783 | 1.15E-06 | 0.001 |
| FCGR1A | 0.515 | 7.46E-05 | 0.013 | 0.331 | 6.29E-04 | 0.043 |
| LILRA5 | 0.481 | 4.90E-04 | 0.025 | 0.452 | 1.87E-04 | 0.024 |
| MAPKAPK3 | 0.222 | 4.12E-06 | 0.003 | 0.117 | 1.38E-04 | 0.020 |
| FNDC4 | 0.192 | 0.001 | 0.039 | 0.229 | 4.41E-04 | 0.035 |
| RPL22L1 | 0.167 | 0.001 | 0.047 | 0.147 | 6.49E-04 | 0.044 |
| TEX30 | 0.156 | 5.06E-04 | 0.026 | 0.166 | 4.07E-06 | 0.004 |
| PSME2 | 0.116 | 5.52E-04 | 0.026 | 0.101 | 4.52E-04 | 0.035 |
| NRDE2 | -0.077 | 0.001 | 0.036 | -0.065 | 8.19E-04 | 0.048 |
| CDC42BPA | -0.097 | 5.91E-05 | 0.012 | -0.084 | 3.18E-04 | 0.031 |
| ITGA1 | -0.124 | 0.001 | 0.037 | -0.111 | 4.47E-04 | 0.035 |
| ZNF827 | -0.144 | 1.55E-04 | 0.017 | -0.157 | 5.64E-06 | 0.005 |
| OXCT1 | -0.146 | 5.08E-04 | 0.026 | -0.096 | 4.28E-04 | 0.035 |
| GXYLT2 | -0.158 | 0.002 | 0.048 | -0.203 | 2.36E-04 | 0.027 |
| C2orf16 | -0.163 | 7.04E-04 | 0.029 | -0.148 | 7.03E-04 | 0.045 |
| RP11-999E24.3 | -0.175 | 0.001 | 0.045 | -0.256 | 6.73E-06 | 0.005 |
| PLCE1 | -0.186 | 7.89E-04 | 0.031 | -0.204 | 6.29E-04 | 0.043 |
| LOX | -0.218 | 5.14E-04 | 0.026 | -0.181 | 8.12E-04 | 0.048 |
| ANK1 | -0.219 | 0.001 | 0.041 | -0.330 | 6.08E-05 | 0.015 |
| LEFTY2 | -0.244 | 0.001 | 0.043 | -0.421 | 1.57E-05 | 0.007 |
| FREM1 | -0.259 | 4.05E-04 | 0.023 | -0.258 | 4.30E-04 | 0.035 |
| MTCO3P12 | -0.784 | 3.53E-06 | 0.003 | -0.789 | 1.06E-07 | 3.12E-04 |

Legend: log2FC denotes log2 fold change; padj denotes adjusted p-value using False Discovery Rate (FDR) method.

### Validation of DEGs in their plasma protein form using UKB

Of 503,317 UKB participants, we studied 52,996 individuals in the plasma proteomics project cohort (15). Of these, 2,918 (5.5%) had DM at baseline. Compared to the non-DM group, individuals with DM were older, more commonly male, had higher body BMI, a greater prevalence of current smoking, elevated systolic blood pressure, and lower diastolic blood pressure. DM was also associated with higher CIMT, PASI, HbA1c, and serum creatinine, but lower serum lipid concentrations. In addition, the DM group exhibited a higher incidence of all future ASCVD events and all-cause mortality (**Supplementary Table 8**)

Of the 22 shared DEGs from the GTEx cohort, 5 are included in the UKB proteomic dataset (ACP5, LEFTY2, LILRA5, OXCT1 and PSME2) and formed the focus of validation analyses. All, except OXCT1, showed statistically significant differences in plasma protein abundance between individuals with and without DM, which were directionally concordant with the transcriptomic data from GTEx (**Supplementary Table 9**). Hence, ACP5, LILRA5, PSME2 and LEFTY2 were prioritised for ongoing validation, of which the former 3 are elevated in the context of DM. Next, we performed age-sex adjusted linear regression to assess the relationships between their abundance and selected cardiovascular risk factors or phenotypes in the whole cohort (**Table 2**). For serum lipids, blood pressure, HbA1c, hsCRP and BMI, all proteins exhibited significant associations (except for LEFTY2 with HbA1C), although not always in a direction expected for adverse cardiovascular phenotype. For PASI, all but LEFTY2 demonstrated significant positive association, whereas for CIMT only LILRA demonstrated significant positive correlation. Notably, when a DM interaction term was added to the models, this identified differential magnitudes of association between many proteins and cardiovascular risk factors/phenotypes in people with versus without DM. Stratified data for groups with and without DM are presented in **Supplementary Table 10** to illustrate these differential associations more fully. These data support the notion that the 4 proteins are broadly associated with cardiovascular metrics, although often differentially in people with DM.

**Table 2:**
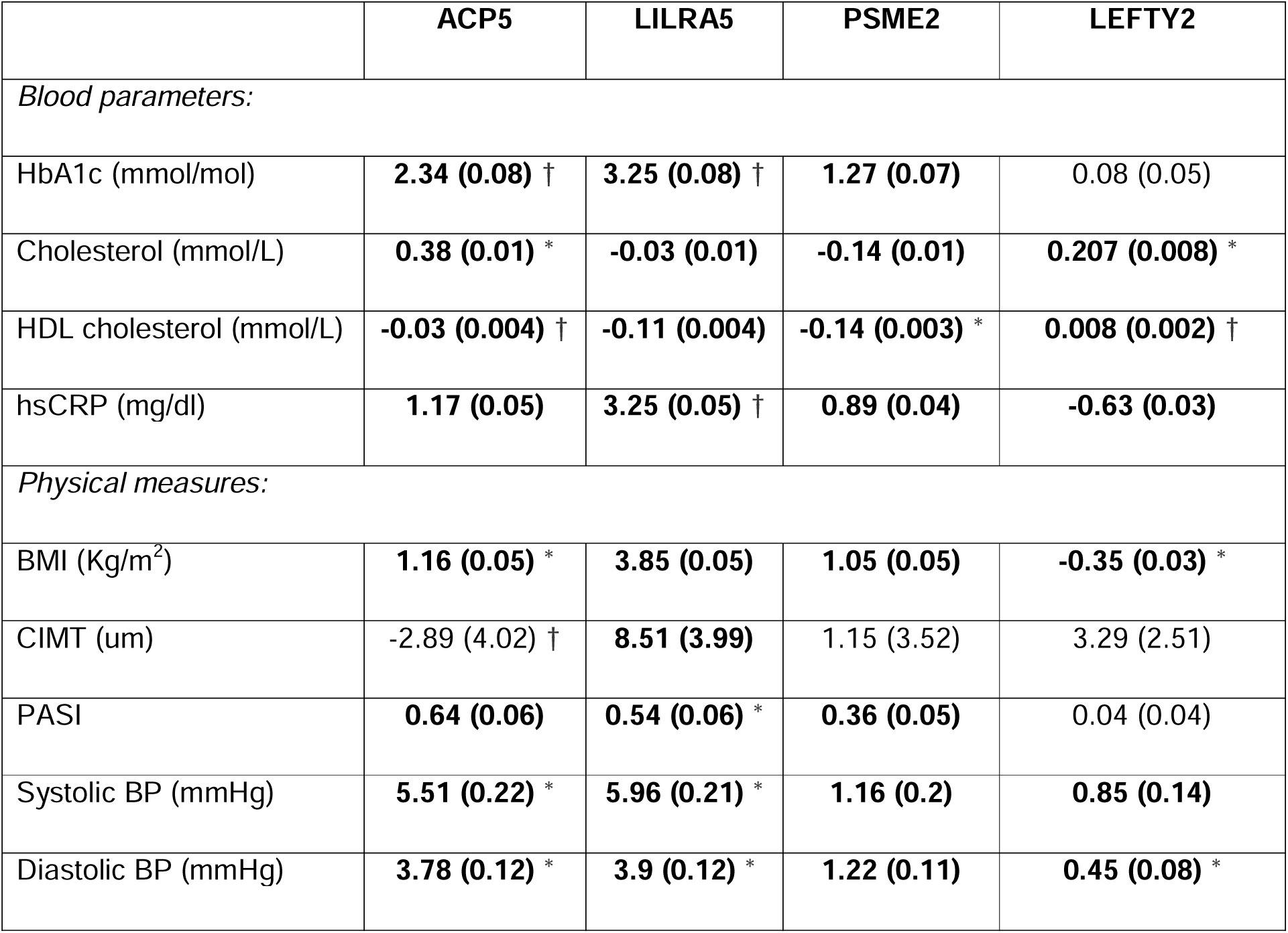
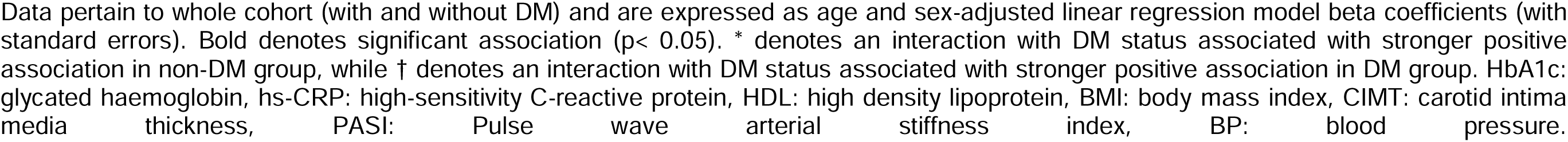
Association *of* ACP5, LILRA5, PSME2 and LEFTY2 *with* cardiovascular risk factors and *phenotypes*.

|  | ACP5 | LILRA5 | PSME2 | LEFTY2 |
| --- | --- | --- | --- | --- |
| *Blood parameters:* |  |  |  |  |
| HbA1c (mmol/mol) | **2.34 (0.08) †** | **3.25 (0.08) †** | **1.27 (0.07)** | 0.08 (0.05) |
| Cholesterol (mmol/L) | **0.38 (0.01) \*** | **-0.03 (0.01)** | **-0.14 (0.01)** | **0.207 (0.008) \*** |
| HDL cholesterol (mmol/L) | **-0.03 (0.004) †** | **-0.11 (0.004)** | **-0.14 (0.003) \*** | **0.008 (0.002) †** |
| hsCRP (mg/dl) | **1.17 (0.05)** | **3.25 (0.05) †** | **0.89 (0.04)** | **-0.63 (0.03)** |
| *Physical measures:* |  |  |  |  |
| BMI (Kg/m<sup>2</sup>) | **1.16 (0.05) \*** | **3.85 (0.05)** | **1.05 (0.05)** | **-0.35 (0.03) \*** |
| CIMT (um) | -2.89 (4.02) † | **8.51 (3.99)** | 1.15 (3.52) | 3.29 (2.51) |
| PASI | **0.64 (0.06)** | **0.54 (0.06) \*** | **0.36 (0.05)** | 0.04 (0.04) |
| Systolic BP (mmHg) | **5.51 (0.22) \*** | **5.96 (0.21) \*** | **1.16 (0.2)** | **0.85 (0.14)** |
| Diastolic BP (mmHg) | **3.78 (0.12) \*** | **3.9 (0.12) \*** | **1.22 (0.11)** | **0.45 (0.08) \*** |

Data pertain to whole cohort (with and without DM) and are expressed as age and sex-adjusted linear regression model beta coefficients (with standard errors). Bold denotes significant association ( $p < 0.05$ ). \* denotes an interaction with DM status associated with stronger positive association in non-DM group, while † denotes an interaction with DM status associated with stronger positive association in DM group. HbA1c: glycated haemoglobin, hs-CRP: high-sensitivity C-reactive protein, HDL: high density lipoprotein, BMI: body mass index, CIMT: carotid intima media thickness, PASI: Pulse wave arterial stiffness index, BP: blood pressure.

To further validate our protein hits, we defined their association with ASCVD events. During 704,470 person-years of follow-up (mean 13.3 years per person), 5,724 participants died (10.8%) of whom 1,219 (2.3%) died from cardiovascular causes. A total of 2,437 participants (4.6%) experienced a non-fatal MI, and 1,423 (2.6%) a non-fatal ischemic stroke. For each protein, participants were stratified into quartiles (Q), with Q1 denoting the lowest plasma protein abundance. Cox proportional hazards regression was used to compare event rates Q4 versus Q1 (**Table 3**), with Kaplan-Meier curves illustrating unadjusted cardiovascular mortality event rates across quartiles (**Figure 2**). This revealed that higher concentrations of ACP5, LILRA5 and PSME2 were associated with greater risk of MI, ischaemic stroke, cardiovascular mortality (except for ACP5) and all-cause mortality. Less consistent associations were noted for LEFTY2, higher concentrations of which were associated with higher risk of cardiovascular mortality only. Inclusion of a DM interaction term in these models revealed similar associations between LILRA5 and all outcomes in the DM and non-DM groups. However, stronger magnitudes of association were noted between ACP5 and PSME2 and some outcomes in the non-DM group. Conversely, stronger magnitudes of association were noted between LEFTY2 and mortality in the DM group. Stratified data for groups with and without DM are presented in **Supplementary Tables 11-12** and to illustrate these differential associations more fully. These data further support the notion that the 4 proteins are broadly associated with cardiovascular metrics, although often differentially according to DM status.

**Figure 2:**
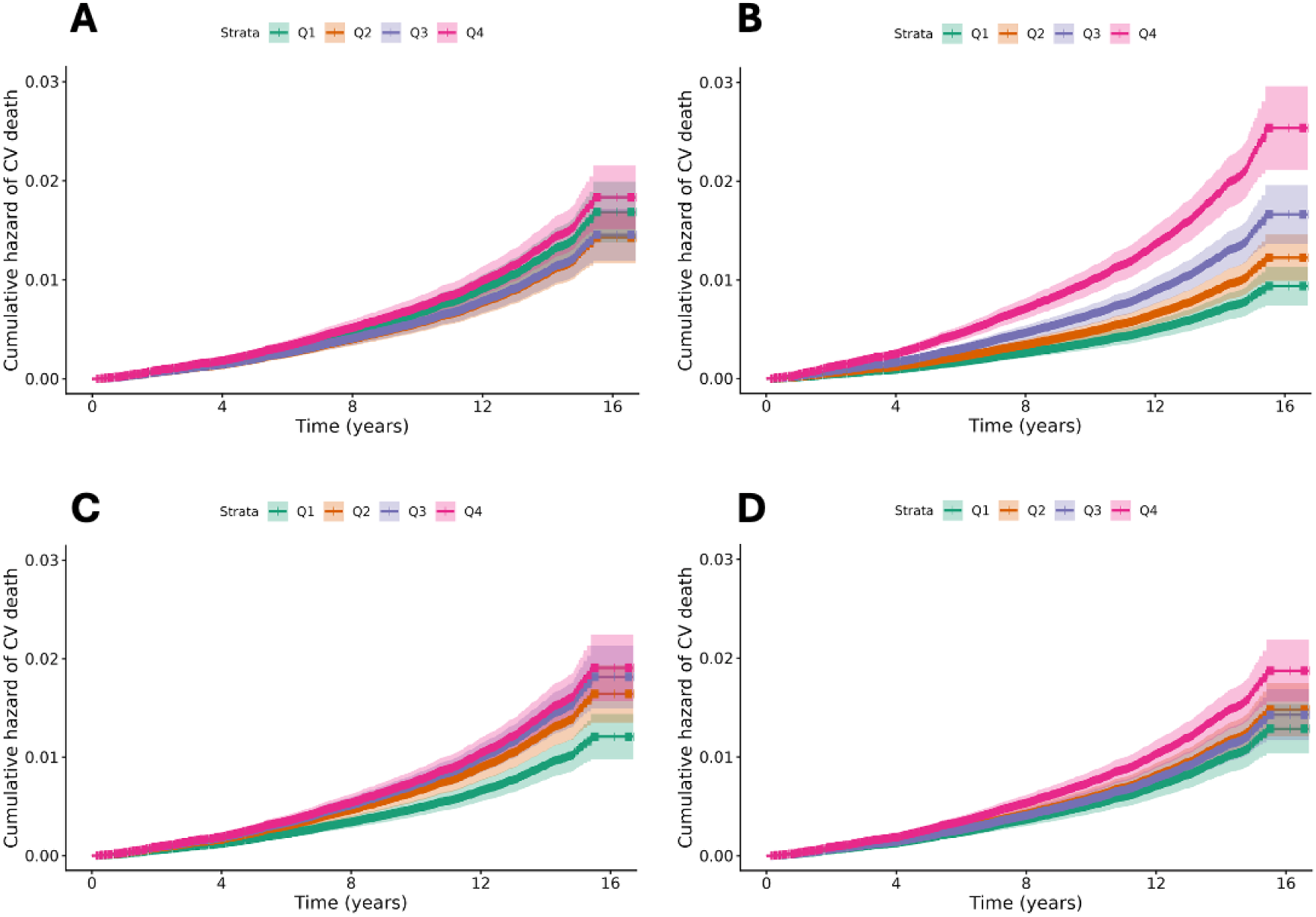
Associations between ACP5, LILRA5, PSME2 and LEFTY2 with cardiovascular mortality. Kaplan-Meier curves illustrating cardiovascular mortality rates according to quartiles (Q) of ACP5 (**A**), LILRA5 (**B**), PSME2 (**C**), and LEFTY2 (**D**). (Q1) represents the lowest protein concentration and Q4 the highest.

**Table 3:** Association *of* ACP5, LILRA5, PSME2 and LEFTY2 *with cardiovascular events*.

|  | **ACP5** | **LILRA5** | **PSME2** | **LEFTY2** |
| --- | --- | --- | --- | --- |
| **All cause death** | **1.33**<br>**(1.23-1.43)**<br>* | **1.83**<br>**(1.69-1.98)** | **1.30**<br>**(1.20-1.40)**<br>* | 1.04<br>(0.96-1.12)<br>† |
| **CV death** | 1.08<br>(0.92-1.27) | **2.70**<br>**(2.25-3.24)** | **1.57**<br>**(1.32-1.88)**<br>* | **1.45**<br>**(1.23-1.72)**<br>† |
| **STEMI** | **1.59**<br>**(1.21-2.09)** | **1.83**<br>**(1.41-2.39)** | **1.57**<br>**(1.19-2.08)** | 0.86<br>(0.66-1.12) |
| **NSTEMI** | **1.51**<br>**(1.28-1.79)** | **1.87**<br>**(1.57-2.23)** | **1.35**<br>**(1.14-1.59)** | 1.04<br>(0.88-1.23) |
| **Ischemic stroke** | **1.31**<br>**(1.11-1.55)**<br>* | **1.54**<br>**(1.30-1.83)** | **1.29**<br>**(1.08-1.53)**<br>* | 1.02<br>(0.86-1.20) |

Age-sex adjusted hazard ratios (and 95% confidence intervals) are presented comparing quartile 4 against quartile 1 of protein concentration. CV: cardiovascular, (N)STEMI: (non-)ST segment elevation myocardial infarction. Bold denotes $p < 0.05$ . \* denotes an interaction with DM status associated with stronger positive association in non-DM group, while † denotes an interaction with DM status associated with stronger positive association in DM group.

### Incremental value of protein hits in ASCVD risk assessment

Finally, we explored whether the 4 proteins (ACP5, LILRA5, PSME2 and LEFTY2) offered incremental insights beyond those offered by clinically applied ASCVD risk prediction tools. To do this, we focused on the SCORE2 and SCORE2-Diabetes models, developed by the European Society of Cardiology (ESC) for use in people with and without DM, respectively. To further ensure that these were calibrated optimally, covariate weightings were refitted to our UKB cohort. Given the proteins’ association with ASCVD events in people with and without DM, we also pooled the 10-year ASCVD risk predictions arising from the non-DM and DM subgroups to increase statistical power, from now on referred to as ‘SCORE2-Pooled’. Amongst our UKB cohort, 35,898 participants had complete data for all criteria required for SCORE2 or SCORE2-Diabetes calculation, along with meeting the criteria for their use (age 40-69 and no prior ASCVD event); all other participants were excluded from the subsequent analyses. Within 10 years of recruitment, 253 (0.7%) died due to CV events and 1358 (3.8%) had non-fatal ASCVD events, with 1490 (4.2%) ASCVD events overall. The cohort was stratified into 34,411 without DM and 1,487 with DM, of which 1,325 (3.8%) and 165 (11.1%) had ASCVD events within 10 years of recruitment, respectively.

To assess the discriminative performance of models, we quantified area under the curve (AUC) from receiver operating characteristic (ROC) curves (**Figure 3A**). The AUC of SCORE2-Pooled (0.722; 95% confidence interval [CI] 0.710-0.735) exhibited a small, but statistically significant (DeLong p=0.04) increase with addition of the 4 proteins (0.725; 95% Cl 0.713-0.738), now referred to as SCORE2-Pooled-4P. To understand how addition of the proteins changed risk estimates for individual participants, we then studied net reclassification index (NRI) using both continuous and categorical approaches. For continuous NRI, we observed that SCORE2-Pooled-4P adjusted classification by 22.3% (95% Cl 17.1-27.7%) with upward reclassification 7.6% and downward classification 14.7% For categorical reclassification, we applied thresholds proposed by ESC to denote boundaries between low, moderate, high and very high risk (at 2%, 10% and 20% predicted risk of 10-year ASCVD event). Addition of the 4 proteins to SCORE2-Pooled led to 5.6% upward and 4.3% downward movement of cases (i.e. a net 1.3% improvement), and to 3.4% upward and 4.3% downward movement of controls (i.e. a net improvement of 0.9%; **Supplementary Table 13**). Finally, we assessed calibration of SCORE2-Pooled and SCORE2-Pooled-4P to define how closely their predicted event rates matched observed event rates and this revealed excellent calibration for both (**Figure 3B**). Overall, these data suggest that ACP5, LILRA5, PSME2 and LEFTY2 provide incremental insights about ASCVD risk, beyond metrics used in routine clinical risk prediction.

**Figure 3:**
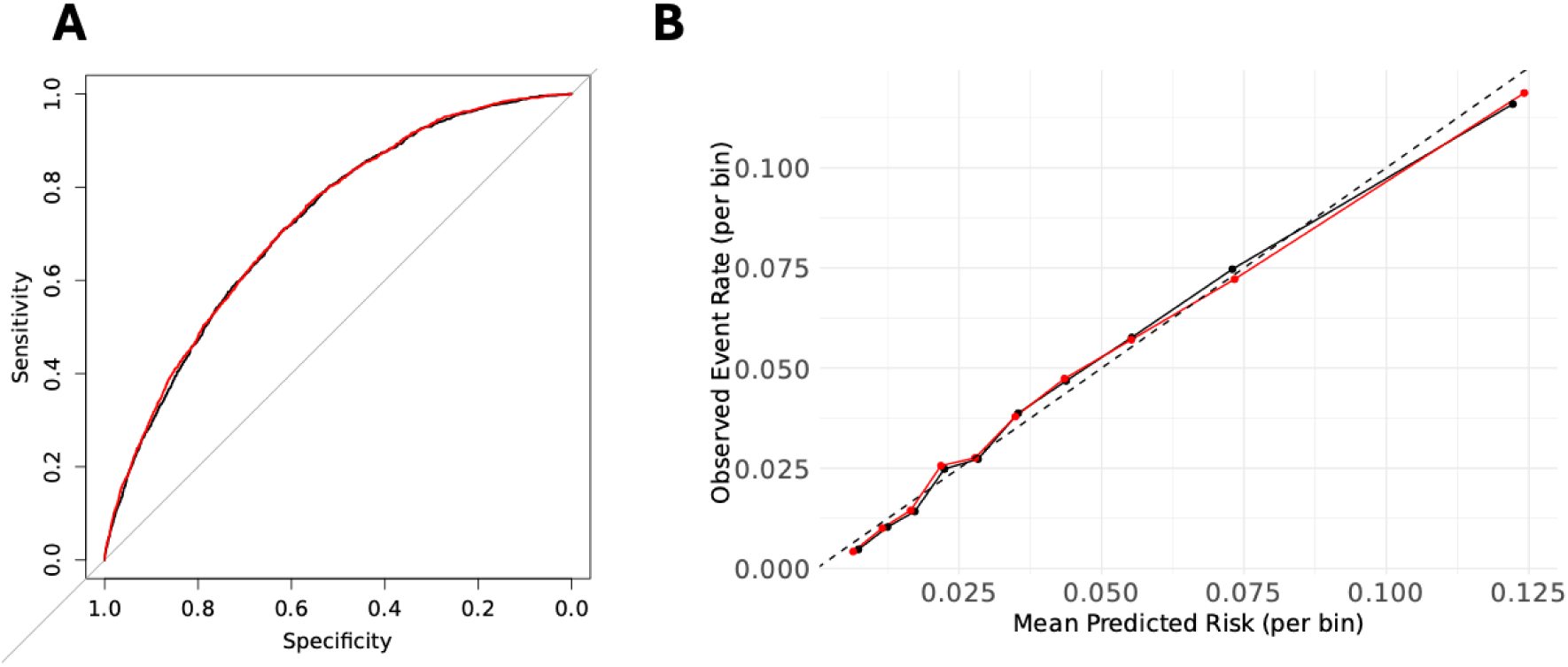
Impact of ACP5, LILRA5, PSME2 and LEFTY on discrimination and calibration of SCORE2-Pooled in predicting atherosclerotic cardiovascular disease events. **A**) Receiver operating characteristic (ROC) curves showing area under curve of both models for SCORE2-Pooled (Black line) and SCORE2-Pooled-4P (which includes ACP5, LILRA5, PSME2 and LEFTY2; Red line). **B**) Calibration plots of the refitted SCORE2-Pooled (Black line) and SCORE2-Pooled-4P (Red line) models. The dashed 45° line indicates the region of perfect calibration.

### Conclusions

Our study includes what we believe is the largest analysis of the arterial transcriptome characterising DM, including two distinct anatomical sites and accounting for multiple donor and technical covariates. This revealed largely non-overlapping signatures in the thoracic aorta and tibial artery, with distinct inflammatory gene sets being enriched in each. However, 22 DEGs were shared in the two arteries, with half of the 8 upregulated genes annotated by the Human Protein Atlas as enriched in macrophages. To validate these shared DEGs, we leveraged the UKB resource to explore hits with available plasma protein data, and corroborated 4 (ACP5, LILRA5, PSME2 and LEFTY2) as being concordantly differentially abundant in people with DM. Notably, these were all associated with ASCVD risk factors, proxies (PASI or CIMT) and incident major clinical events. Moreover, they improved the performance of the European Society of Cardiology-advocated ASCVD risk model SCORE2/SCORE2-Diabetes. Hence, molecular characterisation of the diabetic arterial wall can lead to insights beyond existing risk factors, which could inform future biomarker and therapeutic development.

Even after accounting for confounding factors, including obesity, people with DM experience at least a 50% greater risk rate of ASCVD events (13), with risks spanning all major arterial beds (7). Multiple imaging studies have shown DM is characterised by more widespread and severe coronary atherosclerosis, including greater arterial calcification and lipid accumulation (4,5). This has been corroborated by histological analysis, which also suggests greater macrophage and T lymphocyte infiltration in the arterial wall (6). Molecular insights have largely come from mouse models and *in vitro* experiments, which have implicated diverse processes such as oxidative stress, inflammation, abnormal metabolism (16–18). Our data complement these by arriving at molecular hits using an unbiased assessment of the human arterial wall, suggesting inflammation as a unifying major theme, although with distinct facets in the aorta and tibial artery. These insights are novel in relation to other published transcriptomic studies which were an order of magnitude smaller in sample size (19,20). Half of the genes we found to be upregulated in both vessels are curated by the Human Protein Atlas as enriched in macrophages, and other work has shown DM substantially perturbs macrophage biology via altered metabolism, oxidative stress and epigenetic modification, amongst others (21–23). This suggests a need for further studies of arterial wall derived immune cells from people with DM.

Regarding our four prioritised molecular hits, one has evidence supporting its role in diabetic vascular disease, whereas the potential role of others is either speculative or unclear. ACP5, also known as tartrate-resistant acid phosphatase, is known to influence tissue calcification and myeloid cell biology (24,25), both of which are highly relevant to diabetic vascular disease. Indeed, recent data in a diabetic rat arterial balloon injury model have implicated ACP5 in the inflammatory response to vascular injury (26). LILRA5 is expressed predominantly by myeloid cells, with its expression regulated by inflammatory cytokines (27); crosslinking of LILRA5 between cells further induces inflammatory cytokine generation, with shedding of soluble LILRA5 proposed to regulate its signalling (28). It has not previously been implicated in atherosclerosis or diabetic vascular disease. PSME2 plays a critical role in MHC-class I antigen processing and its expression is augmented by interferon-gamma (29). Whilst it has not been studied in atherosclerosis or diabetic vascular disease, many studies suggest a role in cancer, where it may act as a marker of M1 macrophage polarisation (30). LEFTY2, which unlike the other hits was downregulated in people with DM, is best known for its role in heterotaxy during embryonic development (31). Whilst roles in DM or arterial disease are unknown, it is a member of the TGF-beta superfamily and has recently been reported to regulate bone remodelling, so a potential role in vascular remodelling calcification is conceivable (32). Overall, these data suggest limited understanding of our hits in diabetic arterial disease, warranting further research.

Many other studies have explored the potential of augmenting clinical ASCVD risk scores, for example with imaging, routine biomarker, or multi-omics data. Perhaps the most relevant example in the context of our study, Royer *et al* used UKB data to explore the incremental value of adding up to 2,919 plasma proteins to SCORE2 (33). They found that these, or 115 proteins selected in a data driven process, or 114 proteins selected in a literature driven process, could improve the discriminative performance, up to a peak AUC of 0.775 versus 0.749 for the refitted SCORE2 model. This illustrates the challenge in augmenting model discrimination even with large volumes of additional data and provides important context for the modest increase we observed. It is also notable that we found a relevant degree of reclassification with addition of our four prioritised hits, suggesting that these could still influence clinician and patient behaviour, assuming further validation studies were supportive. However, we believe that a greater value in this analysis is in demonstrating that our hits inform about biology that is not accounted for by traditional risk factors (and our responses to these). This implies that further research to explore their meaning could inform novel therapeutic avenues.

It is also important to address the limitations of our data and analytical approach. First, our data cannot define causality and so additional research using direct manipulation of molecular targets, such as mouse models, and/or causal inference approaches, such as Mendelian Randomisation, will be required to establish such relationships. Second, it must be noted that the GTEx cohort is limited by its focus on post-mortem tissue collection and limited clinical metadata, whilst the UKB cohort is known to be healthier than the wider UK population (34). Despite these issues, it is reassuring that most signals we derived from GTEx were supported by analysis in UKB, although further validation would be important before using the data in clinical practice. Third, our analytical pipeline only allows gene hits to be explored where present in the Olink 3072 plasma proteomics panel used by UKB. This means that 17 hits were not taken on to validation and other approaches will be needed to explore these, such as spatial quantification of gene or protein expression in the arterial wall, which could also aid the assessment of the four prioritised hits. Finally, it is beyond the scope of this project to pursue the hits detected only in the thoracic aorta or tibial artery, although these are also likely to be interesting in defining less generalised impacts of DM in these important vessels.

In summary, we have shown that DM is associated with altered gene expression in the arterial wall, which is largely non-overlapping between the thoracic aorta and tibial artery, although both are over-represented by inflammatory gene sets. Within the commonly dysregulated genes in both vessels, we validated ACP5, LEFTY2, LILRA5 and PSME2 in their plasma protein form and showed that these add valve to a clinical ASCVD risk prediction model. These hits may inform about the underlying biology of diabetic macrovascular disease and warrant further exploration to guide fundamental understanding and address therapeutic potential.

## Acknowledgements

This research has been conducted using the UK Biobank Resource under Application Number 528243. This work uses data provided by patients and collected by the NHS as part of their care and support. This research used data assets made available by National Safe Haven as part of the Data and Connectivity National Core Study, led by Health Data Research UK in partnership with the Office for National Statistics and funded by UK Research and Innovation (research which commenced between 1st October 2020 – 31st March 2021 grant ref MC_PC_20029; 1st April 2021 -30th September 2022 grant ref MC_PC_20058).

## Data availability

The data underlying this article were accessed from the GTEx consortium (https://gtexportal.org) and UK Biobank (https://www.ukbiobank.ac.uk) and are available to other scientists after application to these organisations.

## References

1. World Heart Federation. World Heart Report 2023: Confronting the World’s Number One Killer. Geneva; 2023.

2. Einarson TR, Acs A, Ludwig C, Panton UH. Prevalence of cardiovascular disease in type 2 diabetes: A systematic literature review of scientific evidence from across the world in 2007-2017. Cardiovasc Diabetol. 2018;17(1):83.

3. Saeedi P, Petersohn I, Salpea P, Malanda B, Karuranga S, Unwin N, et al. Global and regional diabetes prevalence estimates for 2019 and projections for 2030 and 2045: Results from the International Diabetes Federation Diabetes Atlas, 9th edition. Diabetes Res Clin Pract. 2019;157:107843.

4. Khazai B, Luo Y, Rosenberg S, Wingrove J, Budoff MJ. Coronary atherosclerotic plaque detected by computed tomographic angiography in subjects with diabetes compared to those without diabetes. PLoS One. 2015 Nov;10(11):e0143187.

5. Kato K, Yonetsu T, Kim SJ, Xing L, Lee H, Mcnulty I, et al. Comparison of nonculprit coronary plaque characteristics between patients with and without diabetes: A 3-vessel optical coherence tomography study. JACC Cardiovasc Interv. 2012 Dec;5(11):1150–8.

6. Yahagi K, Kolodgie FD, Lutter C, Mori H, Romero ME, Finn A V., et al. Pathology of human coronary and carotid artery atherosclerosis and vascular calcification in diabetes mellitus. Arterioscler Thromb Vasc Biol. 2017 Feb;37(2):191–204.

7. Zhao Y, Evans MA, Allison MA, Bertoni AG, Budoff MJ, Criqui MH, et al. Multisite atherosclerosis in subjects with metabolic syndrome and diabetes and relation to cardiovascular events: The Multi-Ethnic Study of Atherosclerosis. Atherosclerosis. 2019 Mar;282:202–9.

8. Lonsdale J, Thomas J, Salvatore M, Phillips R, Lo E, Shad S, et al. The Genotype-Tissue Expression (GTEx) project. Nat Genet. 2013 May;45:580–5.

9. Carithers LJ, Ardlie K, Barcus M, Branton PA, Britton A, Buia SA, et al. A Novel Approach to High-Quality Postmortem Tissue Procurement: The GTEx Project. Biopreserv Biobank. 2015 Oct 1;13(5):311–7.

10. Love MI, Huber W, Anders S. Moderated estimation of fold change and dispersion for RNA-seq data with DESeq2. Genome Biol. 2014 Dec 5;15(12):550.

11. Conning-Rowland MS, Giannoudi M, Drozd M, Brown OI, Yuldasheva NY, Cheng CW, et al. The diabetic myocardial transcriptome reveals Erbb3 and Hspa2 as a novel biomarkers of incident heart failure. Cardiovasc Res. 2024 Dec 4;120(15):1898–906.

12. Ashburner M, Ball CA, Blake JA, Botstein D, Butler H, Cherry JM, et al. Gene Ontology: tool for the unification of biology. The Gene Ontology Consortium. Nat Genet. 2000 May;25(1):25–9.

13. Brown OI, Drozd M, McGowan H, Giannoudi M, Conning-Rowland M, Gierula J, et al. Relationship Among Diabetes, Obesity, and Cardiovascular Disease Phenotypes: A UK Biobank Cohort Study. Diabetes Care. 2023 Aug 1;46(8):1531–40.

14. SCORE2-Diabetes Working Group and the ESC Cardiovascular Risk Collaboration. SCORE2-Diabetes: 10-year cardiovascular risk estimation in type 2 diabetes in Europe. Eur Heart J. 2023 Jul 21;44(28):2557–9.

15. Sun BB, Chiou J, Traylor M, Benner C, Hsu YH, Richardson TG, et al. Plasma proteomic associations with genetics and health in the UK Biobank. Nature. 2023 Oct 12;622(7982):329–38.

16. Khan AW, Jandeleit-Dahm KAM. Atherosclerosis in diabetes mellitus: novel mechanisms and mechanism-based therapeutic approaches. Nat Rev Cardiol. 2025 Jul 1;22(7):482–96.

17. Shah MS, Brownlee M. Molecular and cellular mechanisms of cardiovascular disorders in diabetes. Circ Res. 2016 May 27;118(11):1808–29.

18. Kanter JE, Bornfeldt KE. Inflammation and diabetes-accelerated atherosclerosis: Myeloid cell mediators. Trends Endocrinol Metab. 2013 Mar;24(3):137–44.

19. Skov V, Knudsen S, Olesen M, Hansen ML, Rasmussen LM. Global gene expression profiling displays a network of dysregulated genes in non-atherosclerotic arterial tissue from patients with type 2 diabetes. Cardiovasc Diabetol. 2012 Feb 17;11(15).

20. Qian Y, Xiong S, Li L, Sun Z, Zhang L, Yuan W, et al. Spatial multiomics atlas reveals smooth muscle phenotypic transformation and metabolic reprogramming in diabetic macroangiopathy. Cardiovasc Diabetol. 2024 Dec 1;23(1):358.

21. Rendra E, Riabov V, Mossel DM, Sevastyanova T, Harmsen MC, Kzhyshkowska J. Reactive oxygen species (ROS) in macrophage activation and function in diabetes. Immunobiology. 2019 Mar 1;224(2):242–53.

22. Edgar L, Akbar N, Braithwaite AT, Krausgruber T, Gallart-Ayala H, Bailey J, et al. Hyperglycemia Induces Trained Immunity in Macrophages and Their Precursors and Promotes Atherosclerosis. Circulation. 2021 Sep 21;144(12):961–82.

23. Drareni K, Gautier JF, Venteclef N, Alzaid F. Transcriptional control of macrophage polarisation in type 2 diabetes. Semin Immunopathol. 2019 Jul 1;41(4):515–29.

24. Blumer MJF, Hausott B, Schwarzer C, Hayman AR, Stempel J, Fritsch H. Role of tartrate-resistant acid phosphatase (TRAP) in long bone development. Mech Dev. 2012 Jul;129(5–8):162–76.

25. Bergwik J, Bhongir RKV, Padra M, Adler A, Olm F, Lång P, et al. Macrophage expressed tartrate-resistant acid phosphatase 5 promotes pulmonary fibrosis progression. Immunology. 2024 Apr 1;171(4):583–94.

26. Zhang J, Yang Z, Liu L, Wu D, Cheng C, Zhu P, et al. DOT1L-mediated H3K79me1 transcriptional activation of Acp5 aggravates inflammatory responses following diabetic vascular injury. Microvasc Res. 2026 Jan 1;163:104882.

27. Mitchell A, Rentero C, Endoh Y, Hsu K, Gaus K, Geczy C, et al. LILRA5 is expressed by synovial tissue macrophages in rheumatoid arthritis, selectively induces pro-inflammatory cytokines and IL-10 and is regulated by TNF-α, IL-10 and IFN-γ. Eur J Immunol. 2008;38(12):3459–73.

28. Fu Z, Rumpret M, Kube-Golovin I, Lyndin M, Solntceva V, Zhao Y, et al. LILRA5 Functions to Induce ROS Production on Innate Immune Cells. Eur J Immunol. 2025 Oct 1;55(10):e70079.

29. Sijts A, Sun Y, Janek K, Kral S, Paschen A, Schadendorf D, et al. The role of the proteasome activator PA28 in MHC class I antigen processing. Mol Immunol. 2002;39(3–4):165–9.

30. Li R, Yan L, Jiu J, Liu H, Li D, Li X, et al. PSME2 offers value as a biomarker of M1 macrophage infiltration in pan-cancer and inhibits osteosarcoma malignant phenotypes. Int J Biol Sci. 2024;20(4):1452–70.

31. Saijoh Y, Adachi H, Sakuma R, Yeo CY, Yashiro K, Watanabe M, et al. Left-right asymmetric expression of lefty2 and nodal is induced by a signaling pathway that includes the transcription factor FAST2. Mol Cell. 2000;5(1):35–47.

32. Kim N, Kim JH, Kim K, Kim I, Seong S. Lefty2 prevents RANKL-induced bone loss by inhibiting osteoclast differentiation. BMB Rep. 2025 Sep 16;59(1):78–83.

33. Royer P, Björnson E, Adiels M, Josefson R, Hagberg E, Gummesson A, et al. Large-scale plasma proteomics in the UK Biobank modestly improves prediction of major cardiovascular events in a population without previous cardiovascular disease. Eur J Prev Cardiol. 2024 Oct 1;31(14):1681–9.

34. Fry A, Littlejohns TJ, Sudlow C, Doherty N, Adamska L, Sprosen T, et al. Comparison of Sociodemographic and Health-Related Characteristics of UK Biobank Participants with Those of the General Population. Am J Epidemiol. 2017 Nov 1;186(9):1026–34.

